# A Multiplexed, Next Generation Sequencing Platform for High-Throughput Detection of SARS-CoV-2

**DOI:** 10.1101/2020.10.15.20212712

**Authors:** Marie-Ming Aynaud, J. Javier Hernandez, Seda Barutcu, Ulrich Braunschweig, Kin Chan, Joel D. Pearson, Daniel Trcka, Suzanna L. Prosser, Jaeyoun Kim, Miriam Barrios-Rodiles, Mark Jen, Siyuan Song, Jess Shen, Christine Bruce, Bryn Hazlett, Susan Poutanen, Liliana Attisano, Rod Bremner, Benjamin J. Blencowe, Tony Mazzulli, Hong Han, Laurence Pelletier, Jeffrey L. Wrana

**Affiliations:** Lunenfeld-Tanenbaum Research Institute, Mount Sinai Hospital, Toronto, Ontario, M5G 1X5, Canada; Department of Microbiology, Mount Sinai Hospital/University Health Network, Toronto, Ontario, M5G 1X5, Canada; Department of Molecular Genetics, Donnelly Centre, University of Toronto, Toronto, Ontario, M5S 3E1, Canada; Donnelly Centre for Cellular and Biomolecular Research, University of Toronto, Toronto, Ontario, M5S 3E1, Canada; Department of Biochemistry, Donnelly Centre for Cellular and Biomolecular Research, University of Toronto, Toronto, Ontario, M5S 3E1, Canada

## Abstract

Population scale sweeps of viral pathogens, such as SARS-CoV-2, that incorporate large numbers of asymptomatic or mild symptom patients present unique challenges for public health agencies trying to manage both travel and local spread. Physical distancing is the current major strategy to suppress spread of the disease, but with enormous socio-economic costs. However, modelling and studies in isolated jurisdictions suggest that active population surveillance through systematic molecular diagnostics, combined with contact tracing and focused quarantining can significantly suppress disease spread^1-3^ and has significantly impacted disease transmission rates, the number of infected people, and prevented saturation of the healthcare system^4-7^. However, reliable systems allowing for parallel testing of 10-100,000’s of patients in larger urban environments have not yet been employed. Here we describe “COVID-19 screening using Systematic Parallel Analysis of RNA coupled to Sequencing” (C19-SPAR-Seq), a scalable, multiplexed, readily automated next generation sequencing (NGS) platform^8^ that is capable of analyzing tens of thousands of COVID-19 patient samples in a single instrument run. To address the strict requirements in clinical diagnostics for control of assay parameters and output, we employed a <u>co</u>ntrol-based <u>P</u>recision-Recall and predictive <u>R</u>eceiver Operator Characteristics (coPR) analysis to assign run-specific quality control metrics. C19-SPAR-Seq coupled to coPR on a trial cohort of over 600 patients performed with a specificity of 100% and sensitivity of 91% on samples with low viral loads and a sensitivity of > 95% on high viral loads associated with disease onset and peak transmissibility. Our study thus establishes the feasibility of employing C19-SPAR-Seq for the large-scale monitoring of SARS-CoV-2 and other pathogens.

## Main

The current gold standard diagnostic for SARS-CoV-2 is Real-Time Quantitative Polymerase Chain Reaction (RT-qPCR), which is not readily adaptable to large scale population testing^9^. To establish a population-scale testing platform we designed a SPAR-Seq multiplex primer mix v1 that targets RNA-dependent RNA Polymerase (*RdRP*), Envelope (*E*), Nucleocapsid (*N*), and two regions of the Spike (*S*) gene that correspond to the receptor binding domain (RBD) and the polybasic cleavage site (PBS) (**Fig. 1a and Supplementary Table 1**). The latter two are SARS-CoV-2-specific regions that capture five key residues necessary for ACE2 receptor binding (*Srbd*) and the furin cleavage site (*Spbs*) that is critical for viral infectivity^10,11^. For quality control, we targeted Peptidylprolyl Isomerase B (*PPIB*). Current standard testing strategies for viral pathogens employ gene-specific primers in “all-in-one” qRT-PCR reactions that could in principle be adapted to incorporate barcodes into gene-specific primers. However, to allow for rapid adaptation to test for novel and multiple pathogens, and/or profiling host responses we used a generic oligo-dT and random hexamer primed reverse transcription step followed by multiplex PCR and barcoding in a rapid, readily automated format we call “COVID-19 screening using Systematic Parallel Analysis of RNA coupled to Sequencing’’ or C19-SPAR-Seq (**Fig. 1b** and **Supplementary Table 1**). Although cost is often cited as a concern for NGS-based testing, our platform is cost effective with retail material costs ranging from USD ∼$9 to $6 for 500 *versus* 10,000 sample batch sizes, respectively (**Supplementary Table 2**).

**Fig. 1:**
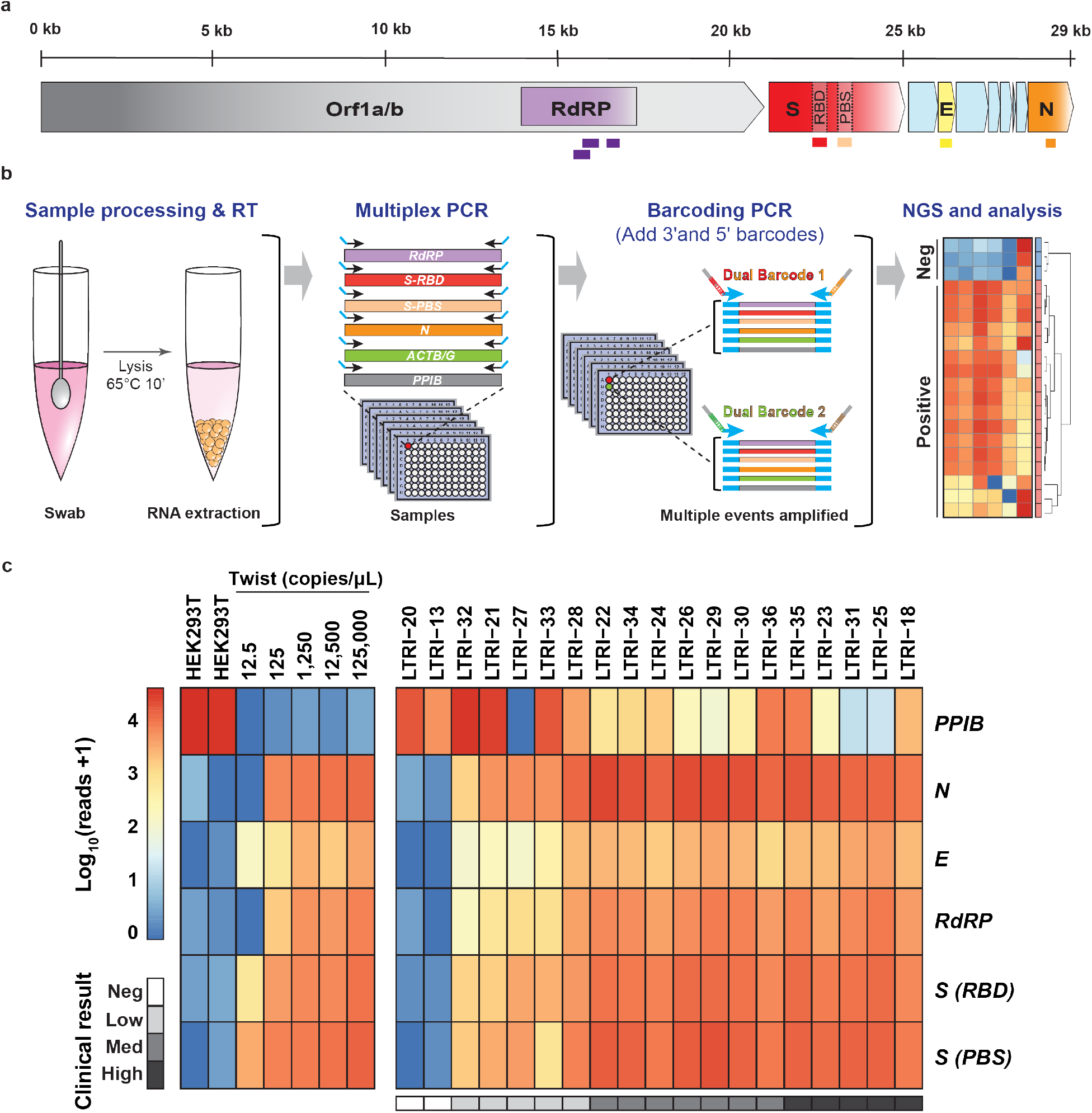
Application of C19-SPAR-Seq to detect SARS-CoV-2. **a**, Schematic representation of the SARS-CoV-2 with the 5 regions targeted for multiplex C19-SPAR-Seq indicated: *RdRP* (purple), *S receptor binding domain* (*Srbd*) (red), *S polybasic cleavage site* (*Spbs*) (light red), *E* (yellow), and *N* (orange). **b**, Schematic of the C19-SPAR-Seq strategy for detecting SARS-CoV-2. cDNA is synthesized using reverse transcriptase (RT) from RNA extracted from clinical samples, subjected to multiplex PCR, then barcoded, pooled and analyzed by next generation sequencing (NGS). **c**, Analysis of archival NASOP swab eluents by C19-SPAR-Seq. A Proof-of-Concept (PoC) cohort (n = 19) was analyzed by C19-SPAR-Seq, and read numbers for each of the indicated amplicons are presented in a heatmap. Control samples (HEK293T, synthetic SARS-CoV-2 RNA) are represented in the left panel, while the right panel shows unsupervised 2D hierarchical clustering of results from negative (blue) and positive (red) patients.

To assess C19-SPAR-Seq performance, we assembled a proof-of-concept (PoC) cohort of 19 archival Nasopharyngeal (NASOP) swab eluents from the Toronto University Health Network-Mount Sinai Hospital clinical diagnostics lab (**Supplementary Table 3**), 17 of which were positive for SARS-CoV-2. Viral load in these archival samples was quantified using the clinically approved TaqMan-based SARS-CoV-2 RT-qPCR detection kit^12^ (‘BGI’), which identified 5 SARS-CoV-2^low^ (Ct > 25), 7 SARS-CoV-2^medium^ (Ct between 20 and 25), and 5 SARS-CoV-2^high^ (Ct < 20) patients (**Supplementary Table 3**). After confirming the efficiency of multiplex v1 primer pairs using a SARS-CoV-2^high^ sample, (LTRI-18, Ct <20; **Extended data Fig. 1**), we performed C19-SPAR-Seq using HEK293T RNA as a negative control (n = 2), and serial dilutions of synthetic SARS-CoV-2 RNA (Twist) as positive controls (n = 5). Pooled sequence data was demultiplexed to individual samples prior to mapping to amplicon sequences. C19-SPAR-Seq was sensitive in detecting as low as 12.5 copies/μL of E, Srbd, and Spbs amplicons from Twist RNA (**Fig. 1c, left panel**). In patient samples, *PPIB* was present in all samples, and all viral targets were robustly detected in high/medium load samples, with reduced detection of *E* and *RdRP* genes in low samples (**Fig. 1c, right panel**).

To establish a diagnostic platform, we performed C19-SPAR-Seq on a larger test development cohort of 24 COVID-19 positive and 88 negative archival patient samples (n = 112; **Supplementary Table 4**). The SARS-CoV-2 RNA standard curve showed a linear relationship between total viral reads and estimated viral copy numbers (**Extended data Fig. 2a**). Negative patient samples had low viral reads (median of 4; range 0-55) compared to positive samples (median of 5,899; range 2-253,956 corresponding to 18 to 705,960 amplicon reads per million reads per sample) (**Fig. 2a**). C19-SPAR-Seq read counts tracked inversely with qRT-PCR Ct values for *RdRP, E* and *N* genes quantified in the diagnostic lab using the Seegene Allplex™ assay^13^ (**Fig. 2b**). Unsupervised clustering showed that the controls performed similarly to the PoC cohort (**Fig. 2c**), as did the positive and negative patient samples, with two exceptions: clinical samples LTRI042 and LTRI050, which displayed background signal, and corresponded to samples with extreme Ct values in only one viral gene (*N* gene, Ct > 38; **Supplementary Table 4**). ROC analysis using total viral reads (**Fig. 2d**) showed excellent performance with an area under the ROC curve (AUC) of 0.969, sensitivity of 92%, specificity of 100%, and overall accuracy of 98%. Thus, C19-SPAR-Seq robustly detects SARS-CoV-2 transcripts, correlates with Ct values from clinical diagnostic tests, and displays excellent performance in distinguishing positive and negative samples.

**Fig. 2:**
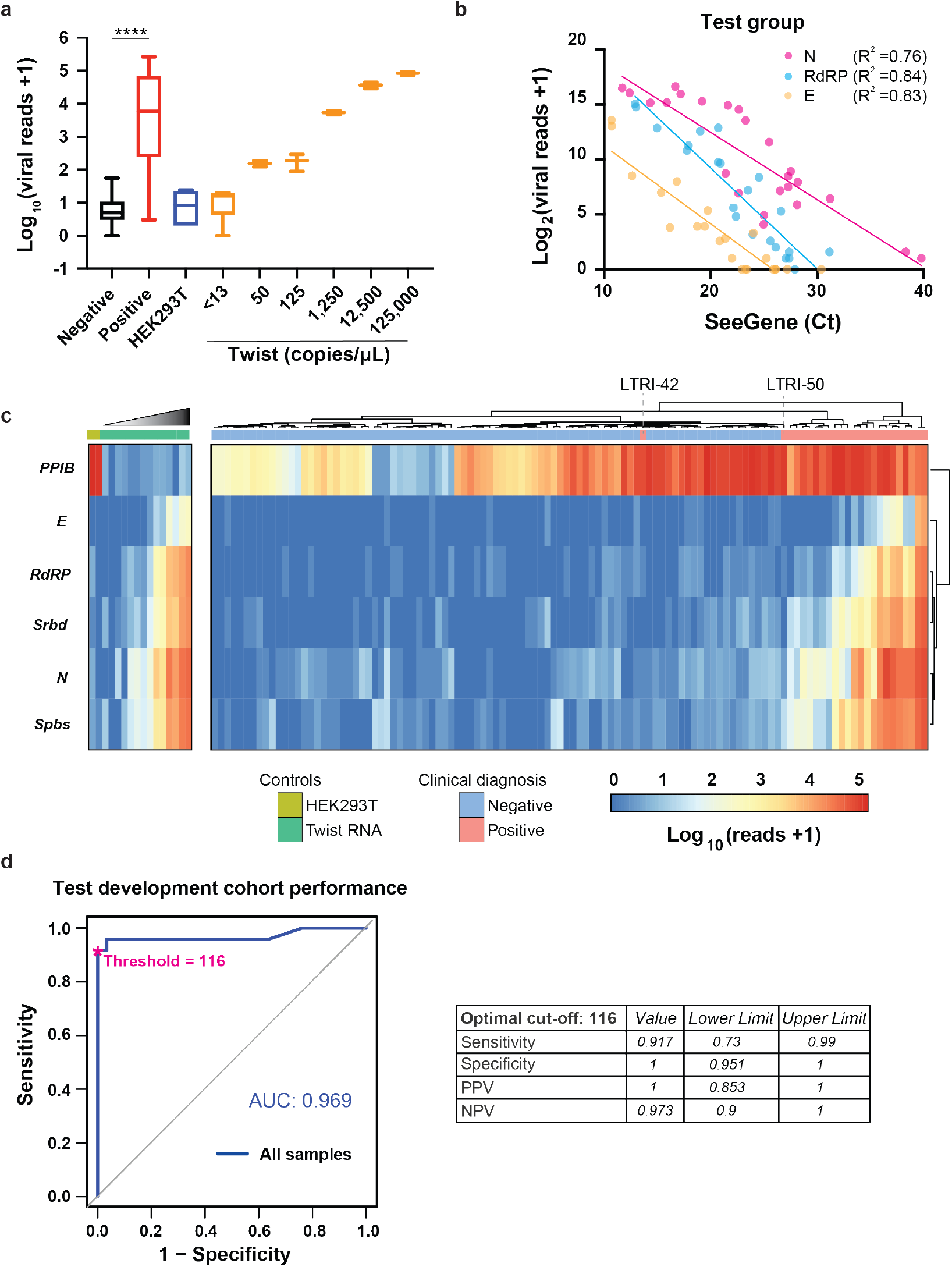
Performance of C19-SPAR-Seq in detecting SARS-CoV-2. **a**, C19-SPAR-Seq of the test development cohort was performed and total viral reads+1 (log_10_) (Y-axis) are plotted for negative (n = 88, black) and positive (n = 24, red) patient samples, HEK293T RNA (n = 6, blue), and the indicated serial dilutions of synthetic SARS-CoV-2 RNA (n=2-6, orange). For each group, the median, lower and upper confidence limit for 95% of the median are plotted. Whiskers are minimum and maximum values. Unpaired *t*-test of negative *versus* positive samples (****: *p* < 0.0001) **b**, C19-SPAR-Seq reads for the indicated gene in each patient sample were compared to Ct values obtained by the clinical diagnostics lab using the ‘Seegene’ Allplex assay. **c**, Heatmap of C19-SPAR-Seq results. Read counts for the indicated target amplicons in control samples (n = 16; left) and patient samples (n = 112; right) are plotted according to the scale, and sample types labelled as indicated. Samples are arranged by hierarchical clustering with euclidean distance indicated by the dendrogram on the top, which readily distinguishes positive from negative samples. **d**, Performance of C19-SPAR-Seq. ROC analysis on patient samples was performed using clinical diagnostic results (Seegene Allplex qRT-PCR assay, **Supplementary Table 3**) and total viral reads for patient samples (n = 112). AUC (area under the curve) scores are indicated on the graph (left), with statistics at the optimal cutoff as indicated (right).

Robust application of C19-SPAR-Seq as a diagnostic tool requires assigning thresholds for both viral RNA detection, as well as host RNA for filtering poor quality samples. In qRT-PCR diagnostics, external validation studies and rigorous standard operating procedures establish pre-defined cutoffs for sample quality and positive *versus* negative assignment (Seegene^13^; BGI^12^). However, in scalable, massively parallel, multiplexed NGS assays, variability in sample numbers and flow cell loading can create run-to-run variations in read numbers, while index-mismatching^14^, as well as trace cross-contamination events can create technical noise that are challenging to control. Furthermore, external validation strategies create a laborious path to adapt and test new multiplex designs to SARS-CoV-2, additional respiratory pathogens, or host responses.

We therefore exploited the throughput of C19-SPAR-Seq to include in every run a training set of large numbers of controls that can be exploited to define cutoffs tailored to each C19-SPAR-Seq run (**Fig. 3a**). To define quality metrics, we computed precision-recall (PR) curves for classifying control samples as either negative (H2O blanks), or positive for any anticipated amplicon (HEK293T for PPIB or synthetic SARS-CoV-2 RNA for viral amplicons) and calculated the highest F1 score, which is the harmonic mean of PR and a common measure of classifier accuracy (**Fig. 3b**). When mapped onto a ROC curve this corresponded to the region closest to perfect sensitivity and specificity (0, 1) (**Extended data Fig. 2b**). To define the threshold for identifying SARS-CoV-2 positive cases, we next analyzed the embedded standard curve of synthetic SARS-CoV-2 RNA. This displays a linear relationship over 4 orders of magnitude and extends to lower limits of detection indistinguishable from background reads from HEK293T cells (**Fig. 2a and Extended data Fig. 2a**), thus allowing us to identify the viral read count in each C19-SPAR-Seq run that most accurately distinguishes positive from negative (**Fig. 3a**). To identify this threshold, we computed PROC01, which optimizes negative predictive value (NPV) and positive predictive value (PPV)^15^ and defined a point (88 viral reads) close to perfect PR (**Fig. 3c**) and sensitivity and specificity on the ROC curve (**Extended data Fig. 2c**). Importantly, these methods control for run-specific variables by employing training sets that are embedded in every C19-SPAR-Seq run.

**Fig. 3:**
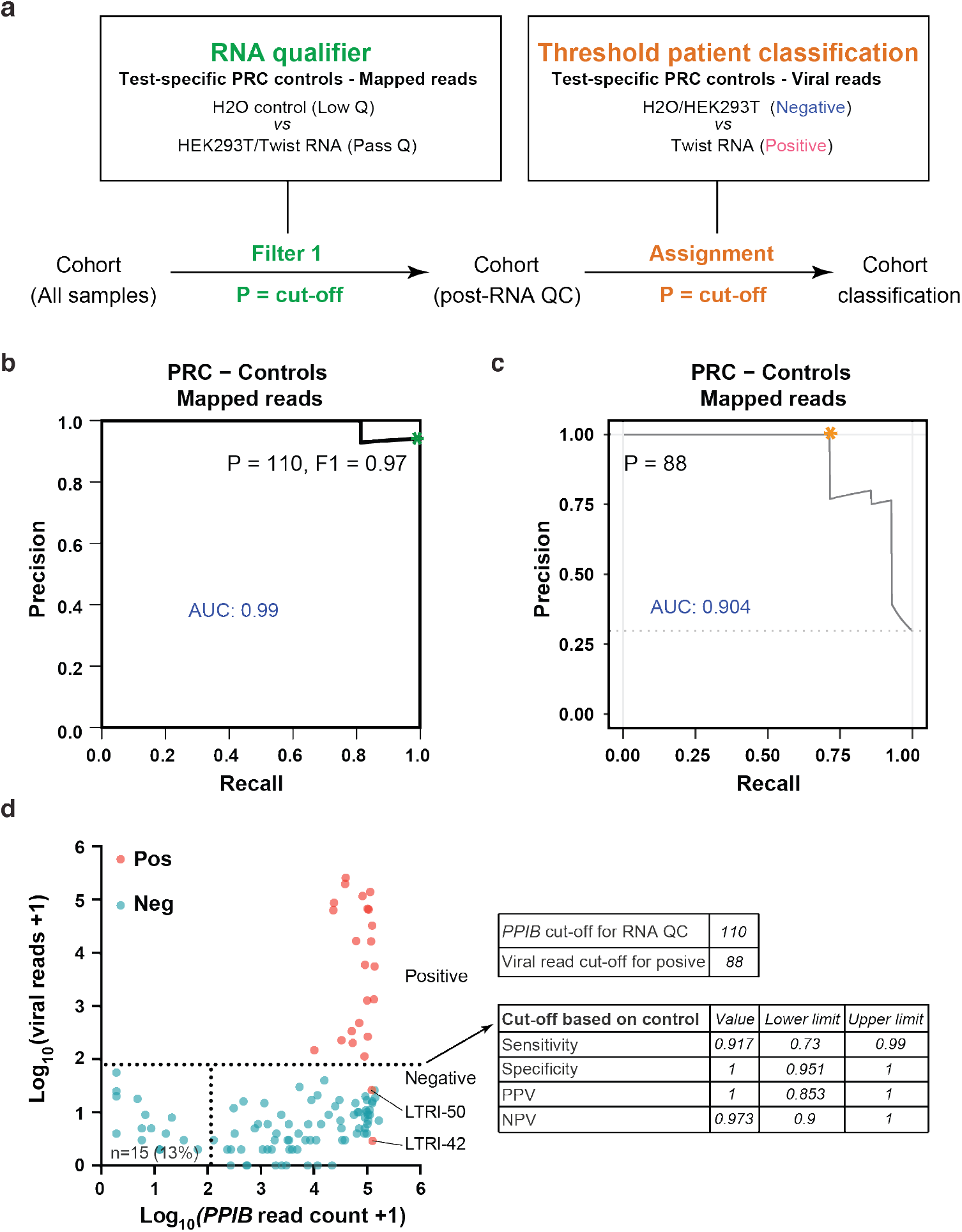
Performance of C19-SPAR-Seq in detecting SARS-CoV-2 using control based classifier. **a**, Schematic of the control based cut-off procedure for RNA quality and viral threshold by coPR analysis. **b**, Thresholding sample quality. coPR analysis on control samples: PRC of control samples for accurate detection of mapped reads are plotted. The optimal precision and recall read cut-off associated (P = 110) with the highest F1 (0.97) score, and AUC (area under the curve) are indicated in the PR plot.. **c**, Threshold for classification of positives in the test cohort. Optimum cut-off for viral threshold is calculated by PROC01 using clinical diagnosis and total viral reads, and plotted on the precision-recall curve. **d**, Threshold assignments for sample quality and classification. Total viral reads +1 (Y-axis) are plotted against *PPIB* reads +1 (X-axis) for positive (red) and negative (blue) patient samples. coPR-based RNA-QC filter and viral read filter are shown as indicated. Assay statistics using coPR thresholding are listed (right).

We next mapped the control-based cutoffs onto patient SPAR-Seq data (**Fig. 3d)**. This showed 15 of these archival samples had low *PPIB* counts that may be due to lost RNA integrity upon repeated freeze-thaw cycles (**Fig. 3d** and **Supplementary Table 4**), a variability we also observed in the PoC cohort (**Fig. 1c**). Of note, C19-SPAR-Seq performance was not affected by filtering poor quality samples (AUC = 0.970; **Extended data Fig. 2d**). Furthermore, using PROC01 thresholding of viral reads identified 22/24 positives with no false positives (**Fig. 3d**). This yielded an overall test performance of 92% sensitivity, 100% specificity, and 98% accuracy (**Fig. 3d** and **Supplementary Table 5)**. This is similar to the observed performance of C19-SPAR-Seq on clinical samples quantified by ROC analysis (**Fig. 2d and Extended data Fig. 2d**, respectively). Thus, an extensive array of internal reference samples is effective as an embedded training set for implementing a <u>co</u>ntrol-based <u>P</u>R/p<u>R</u>OC classifier (coPR) tailored to each C19-SPAR-Seq run.

To validate our C19-SPAR-Seq platform we established a pilot cohort of 378 samples that contains 89 positive samples collected in May of 2020. We first screened for positivity using the clinically approved BGI SARS-CoV-2 kit^12^ which showed 52 samples were positive with > 4 viral copies/μL (**Supplementary Table 6**,**9**). Of the 37 failed samples, 86% had very low viral RNA (only 1 or 2 of the 3 genes detected and/or Ct > 35 on the ‘Seegene’ platform) that may have lost integrity upon storage. Indeed, comparison of Ct values for RdRP detection showed an overall increase of 4 cycles in these archived samples (**Extended data Fig. 3a**), despite the high sensitivity of the BGI platform^16^. The cohort also contained 289 negative samples collected prior to Ontario’s^17^ first confirmed COVID-19 positive case in January 20, 2020, and 1 negative sample collected in May, 2020 (**Supplementary Table 6, Table 1**), and included broncho-alveolar lavages (BALs) and NASOP swabs. Surprisingly, the detection of human RNA dropped substantially to a median of 29 (range 0-41,874), compared to 15,058 (range 2-170,870) in the original test cohort. coPR filtering (**Extended data Fig. 3b**), marked 50% of samples as inconclusive compared to 13% in the test cohort (**Extended Data Fig. 3c**), despite similar distribution of raw reads per sample (**Extended data Fig. 3d**), while mapping rates in the PoC, test and pilot cohorts, progressively declined to as low as 0.1% (**Extended Data Fig. 3e)**. To understand this collapse we analyzed unmapped reads and found that > 90% were consumed by non-specific amplification products (NSAs; **Extended Data Fig. 4a**) that comprised complex chimeric combinations of many viral and human primers (**Extended Data Fig. 4a, b**). For example, RdRP and PPIB contributed to 4 of the top 5 NSAs (NSA1-4), and 2 had a spurious sequence (NSA4,5). Indeed, analysis of C19-SPAR-Seq PoC, test and pilot libraries using a Bioanalyzer, showed that as cohort size and number of negatives increased, NSAs were more apparent, and dominated the pilot library **(Extended data Fig. 4c)**. This suggests that NSAs, enriched in negative samples (3.7-fold increase in the pilot cohort), clog the NGS pipeline as sample numbers rise (**Supplementary Table 9**). This has serious implications for deploying an NGS platform in a population-scale COVID-19 surveillance strategy and highlights the importance of using large-scale cohorts during the development of multiplex testing platforms.

SARS-CoV-2 RNA concentration spans a large dynamic range, such that spike-in mutant amplicons which have been suggested to improve performance of NGS-based strategies^18^ might interfere with detection of COVID-19 positive cases with low viral reads. Therefore, we instead used our NSA data to create multiplex panel v2.0 (see Methods) that removed primers yielding NSAs by targeting a distinct region of *RdRP*, removing *E* and *N* genes, and switching to primers that amplify intron spanning regions of the *ACTB* and *ACTG* genes (**Supplementary Table 1 and Extended data Fig. 1**). We extended the pilot cohort to 663 samples that included 98 confirmed positives and performed C19-SPAR-Seq, which showed targeted amplicons were the predominant product generated by multiplex panel v2.0 (**Extended data Fig. 5a**), and mapping percentages were restored to test cohort levels (**Extended data Fig. 5b**). Total viral read distributions for multiplex panel v2.0 showed good separation in clinically positive samples (**Fig. 4a** and **Extended data Fig. 5c**), while applying coPR thresholding (**Extended data Fig. 5d**) identified 121 samples as inconclusive (**Fig. 4a**), all of which were older, pre-COVID19 material. Of these, 112 were BALs (40% of all BALs), 1 was a bronchial wash (BMSH), and only 8 were NASOPs (1.8% of all NASOPs) (**Supplementary Table 7**). Furthermore, analysis of 10 BAL samples below the QC threshold revealed little or no RNA, contrasting BALs with moderate levels of *ACTB/G* transcripts (representative examples in **Extended data Fig. 6a**), and BAL *ACTB/G* read distributions were much lower than NASOPs (**Extended data Fig. 6b**). This suggests that archival BALs suffered from substantive sample degradation and also highlights how coPR-based thresholding successfully identifies poor quality samples and readily adapts to the use of distinct primer sets.

**Fig. 4:**
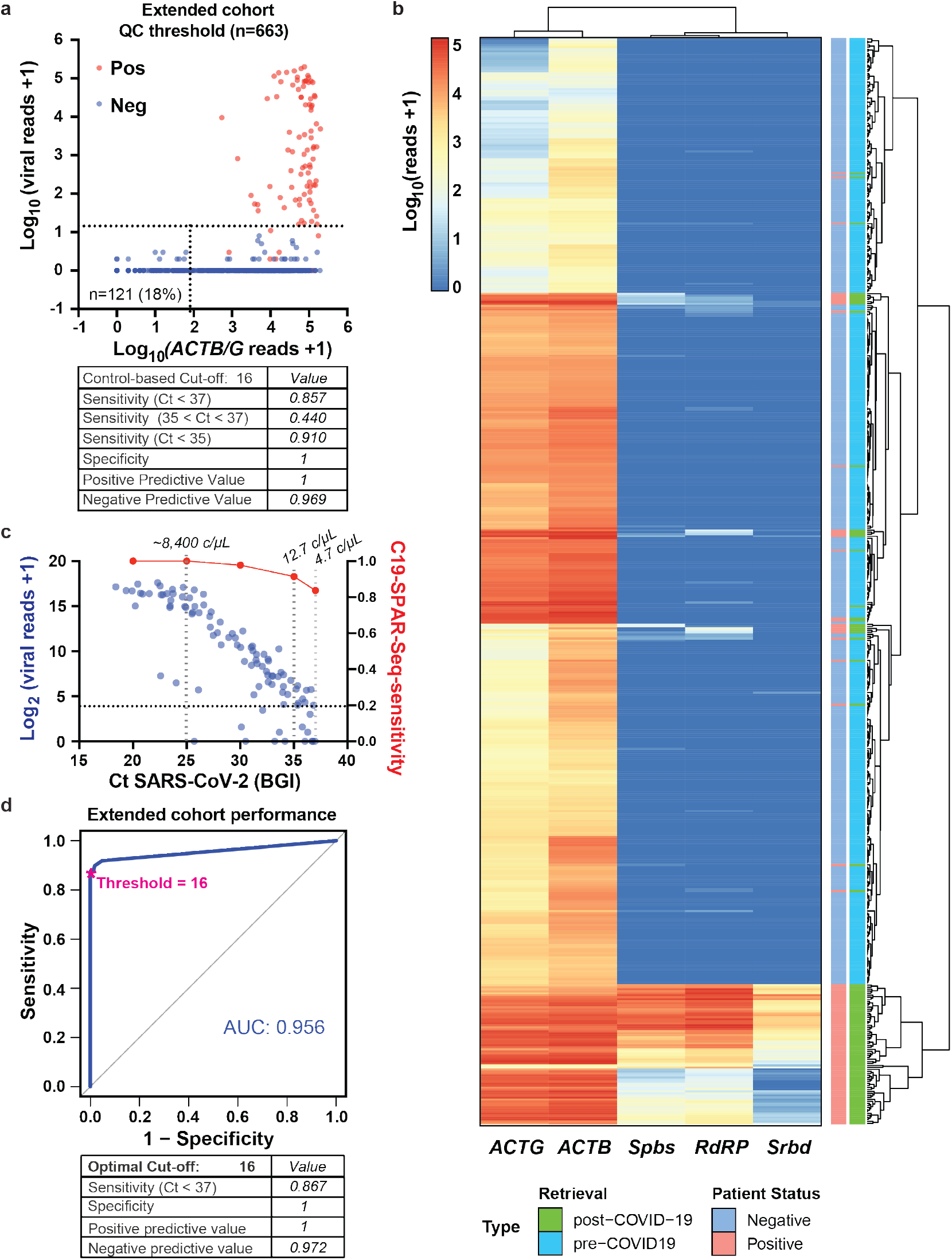
C19-SPAR-Seq of a large patient cohort. **a**, C19-SPAR-Seq on an extended patient cohort. coPR thresholds for sample quality and classification of a 663 patient cohort of negative (blue) and positive (red) specimens are shown as in Fig. 2a. Performance metrics for sample classification according to coPR thresholding are shown in the table. **b**, Heatmap of C19-SPAR-Seq results. Read counts for the indicated target amplicons in the filtered set of samples (n = 542) are plotted according to the scale, and sample types labelled as indicated. Samples are arranged by hierarchical clustering with euclidean distance indicated by the dendrogram on the right. **c**, Scatter plot of total viral reads+1 (left Y-axis, blue) *versus* Ct values of positive samples (n = 98, BGI) (X-axis). C19-SPAR-seq sensitivity at the indicated Ct values is overlaid (right Y-axis, red). Gray dashed lines indicate average copies/μL (c/μL) **d**, ROC curve analysis. ROC curves were processed on filtered samples (n = 542). AUC scores are indicated for filtered samples (blue; left) with corresponding performance statistics for the optimal cut-off indicated below.

Next, we analyzed viral reads, which had a broad range in positive samples (median = 680.5 reads per sample, range 0-200,850; **Fig. 4a** and **Extended data Fig. 5c**). Two-dimensional clustering showed background SARS-CoV-2 products in negative samples were low to undetectable, and *ACTB* typically yielded higher reads than *ACTG*, likely reflecting their differential expression (**Fig. 4b**). Positive samples were generally well separated, although some distinct clusters with lower SARS-CoV-2 reads were apparent (**Fig. 4b** and **Extended data Fig. 5e**). Indeed, total read distributions in positive samples displayed biphasic distribution (**Extended data Fig. 5e**), similar to observations made from RT-qPCR analyses of ∼4000 positive patients^19^. Since the early rapid increase in SARS-CoV-2 viral load at symptom onset is followed by a long tail of low viral load during recovery^20,21^, this biphasic distribution could reflect patients in distinct phases of the disease. We also assessed viral amplicon sequences which matched the SARS-CoV-2 reference (MN908947.3^22^) and found no variants (data not shown). Since neutralizing antibodies are generally thought to target the critical region of the RBD analyzed here^17^, these results suggest the emergence of variant strains that might bypass acquired immunity is not a major feature of SARS-CoV-2. In addition, this supports the notion that biologic therapies targeting the RBD may show broad activity in the population.

We next compared performance of multiplex panel v2.0 to v1.0 using the embedded controls, which showed similar performance (AUC = 0.90, **Extended data Fig. 5f** *versus* 0.92, **Extended data Fig. 2c**, respectively), with coPR yielding an optimal read cutoff of ≤ 16 total viral reads (**Extended data Fig. 5f**) that corresponded to a technical sensitivity of 3 viral copies/μL **(Extended data Fig. 6c)**. coPR thus identified 82 positive samples (**Fig. 4a** and **Supplementary Table 7**) all of which were BGI-confirmed cases to give an overall sensitivity of 86%, specificity of 100%, and accuracy of 97% (**Supplementary Table 8**). Importantly, total viral reads tracked with BGI Ct values (**Fig. 4c**), and for samples with Ct < 35 (corresponding to ∼12 viral copies/μL of specimen), sensitivity was similar to the test cohort at 91%. However, for samples with Ct between 35-37 (4-12 viral copies/μL) sensitivity dropped markedly to 44% (**Supplementary Table 8**), whereas at higher viral loads (Ct = 25 or ∼8,400 viral copies/μL) sensitivity rose to 100% (**Fig. 4c**). ROC analysis of actual C19-SAR-Seq performance yielded an AUC of 0.96, sensitivity of 87% and specificity of 100%, similar to coPR (**Fig. 4d**), while individual amplicons each underperformed total viral reads (AUC: 0.85-0.94; **Extended data Fig. 6d**). Our cohort was biased for samples with very low to low viral loads, which represents a small portion of the COVID-19 population^19^. Therefore, we mapped our sensitivity data onto the population distribution of viral load data^19^, which showed C19-SPAR-Seq sensitivity of ∼97% for patients displaying > 10,000 viral copies/mL (**Extended data Fig. 6e**), which encompasses ∼90% of the positive population. Altogether, these results demonstrate that at high patient sample loads of predominantly negative samples, C19-SPAR-Seq using coPR displays 100% specificity and > 95% sensitivity at viral loads typically observed in populations.

Systematic population-scale testing has been identified as an important tool in managing pandemics such as SARS-CoV-2, where large numbers of infected individuals display mild or no symptoms yet transmit disease. The scalable throughput of C19-SPAR-Seq combined with its excellent sensitivity and specificity at reasonable cost make it well-suited for this role. Data generated by large-scale routine testing of local and larger communities, with different interaction levels would provide valuable epidemiologic information on mechanisms of viral transmission, particularly when coupled to multiplex panels targeting regions of sequence variance currently development. In addition, the C19-SPAR-Seq platform can be readily adapted to incorporate panels tracking multiple pathogens, as well as host responses. C19-SPAR-Seq quantitation would also facilitate real-time tracking of viral load dynamics in populations that may be associated with COVID-19 expansion or resolution phases^20^. Although C19-SPAR-Seq is dependent on centralized regional facilities, it is readily coupled to saliva-based, at-home collection that exploits extensive transport infrastructure and industrialized sample processing to enable frequent widespread testing.

## Methods

### Samples collection

Patient samples (**Supplementary Table 2, 3, 5**) were obtained from the Department of Microbiology at Mount Sinai Hospital under MSH REB Study #20-0078-E, ‘Use of known COVID-19 status tissue samples for development and validation of novel detection methodologies’.

### Total RNA extraction

Total RNA was extracted by using the Total RNA extraction kit (Norgen Biotek kit, Cat. #7200) for the samples in Supplementary Table S2 following the manufacturers guidelines. For all other samples (**Supplementary Table 3, 5**), total RNA was purified in 96 well plates using RNAclean XP beads (Beckman, A66514) and a customized protocol. Briefly, 75.25 μL of patient swabs in transfer buffer were mixed with 14.5 μL of 10X SDS lysis buffer (1% SDS, 10mM EDTA), 48 μL of 6M GuHCl, and 7.25 μL proteinase K (20 mg/mL, ThermoFisher, 4333793), incubated at room temperature for 10’ and heated at 65°C for 10’ prior to the addition of 145 μL of beads. Beads were washed twice in 70% ethanol using a magnetic stand and then RNA eluted into 30 μL Resuspension buffer supplied with the kit. RNA quality was assessed using a Bioanalyzer (5200 Agilent Fragment Analyzer). HEK293T RNA was extracted using the Total RNA extraction kit (Qiagen). Synthetic Twist SARS-CoV-2 RNA (Twist Bioscience #102024 - MN908947.3) was used as positive control.

### Reverse Transcription (RT)

Total RNA was reverse transcribed using SuperScript™ III Reverse Transcriptase (Invitrogen) in 5X First-Strand Buffer containing DTT, a custom mix of Oligo-dT (Sigma) and Hexamer random primers (Sigma), dNTPs (Genedirex) and Ribolock RNase inhibitor (ThermoScientific). We followed the manufacturer’s protocol. Each reaction included: 0.5 μL Oligo-dT, 0.5 μL hexamers, 4 μL purified Total RNA, 1 μL dNTP (2.5 mM each dATP, dGTP, dCTP and dTTP), *quantum satis* (*qs*) 13 μL RNase/DNase free water. Samples were incubated at 65°C for 5’, and then placed on ice for at least for 1’. The following was added to each reaction: 4 μl 5X First-Strand Buffer, 1 μl 0.1 M DTT, 1 μl Ribolock RNase Inhibitor, 1 μl of SuperScript™ III RT (200 units/μl) and then mixed by gently pipetting. Samples were incubated at 25°C for 5’, 50°C for 60’, 70°C for 15’ and then store at 4°C.

### TaqMan-based RT-qPCR detection

A Real-Time Fluorescent RT-PCR kit from ‘BGI’ was used according to manufacturer’s instructions. Experiments were carried out in a 10μl reaction volume in 384-well plates, using 3 μl of sample (LTRI patient samples or Twist RNA), and were analyzed using a Bio-Rad CFX384 detection system (**Supplementary Tables 3**,**6**,**7**). Real-time Fluorescent RT-PCR results from ‘Seegene’ assay were provided by the Department of Microbiology diagnostic lab at Mount Sinai Hospital (**Supplementary Tables 3**,**4**,**6**,**7**).

### C19-SPAR-Seq primer design and optimization

Optimized multiplex PCR primers for SARS-CoV-2 (*N, S, E* and *RdRP*) and human genes (*PPIB and ACTB/G*) were designed using the SPAR-Seq pipeline^8^, with amplicon size > 100 bases (**see Supplementary Table 1**). For *S* gene, two regions were monitored the *S receptor binding domain* (*Srbd*), and *S polybasic cleavage site* (*Spbs*). The Universal adapter sequences used for sequencing were F: 5’-acactctttccctacacgacgctcttccgatct and R: 5’-gtgactggagttcagacgtgtgctcttccgatct). Primers were optimized to avoid primer-dimer and non-specific multiplex amplification. To assess the primers sensitivity and specificity, we performed qPCR (SYBR green master mix, BioApplied) on cDNA prepared from patient samples. Each primer was used at 0.1μM in qPCR reaction run on 384 well plates using Biorad CFX 384 detection system. The thermal cycling conditions were as follows: one cycle at 95°C for 2’, and then 40 cycles of 95°C for 15’’, 60°C for 15’’, 72°C for 20’’, followed by a final melting curve step.

### Multiplexing PCR

The multiplex PCR reaction was carried out using Phusion polymerase (ThermoFisher). The manufacturer’s recommended protocol was followed with the following primer concentrations: all primers (*N, Spoly, Srbd, E, RdRP*, and *PPIB*) were at 0.1 μM for the PoC cohort (**Supplementary Table 3**), SARS-CoV-2 primers (*N, Spoly, Srbd, E* and *RdRP*) were at 0.05 μM, and *PPIB* primer was at 0.1μM for the test and pilot cohort (**Supplementary Table 4, 6**), all primers (*Spoly, Srbd, RdRP* and *ACTB/G*) were at 0.05 μM for the extended cohort (**Supplementary Table 7**). For each reaction: 5 μL 5X Phusion buffer, 0.5 μL dNTP (2.5 mM each dATP, dGTP, dCTP and dTTP), 0.25 μL for each human primers (10 μM), 0.125 μL for each SARS-CoV2 primers (10 μM), 2 μL of cDNA, 0.25 μL Phusion Hot start polymerase, *qs* 25 μL RNase/DNase free water. The thermal cycling conditions were as follows: one cycle at 98°C for 2’, and 30 cycles of 98°C for 15’’, 60°C for 15’’, 72°C for 20’’, and a final extension step at 72°C for 5’ and then stored at 4°C for the PoC and extended cohorts (**Supplementary Table 3, 7**), one cycle at 98°C for 2’, and 35 cycles of 98°C for 15’’, 60°C for 15’’, 72°C for 20’’, and a final extension step at 72°C for 5’ and then stored at 4°C for the test and pilot cohorts (**Supplementary Table 4, 6**).

### Barcoding PCR

For multiplex barcode sequencing, dual-index barcodes were used^8^. The second PCR reaction on multiplex PCR was performed using Phusion polymerase (ThermoFisher). For each reaction: 4 μL 5X Phusion buffer, 0.4 μL dNTP (2.5 mM each dATP, dGTP, dCTP and dTTP), 2 μL Barcoding primers F+R (pre-mix), 4 μL of multiplex PCR reaction, 0.2 μL Phusion polymerase, *qs* 20μL RNase/DNase free water. The thermal cycling conditions were as follows: one cycle at 98°C for 30’’, and 15 cycles of 98°C for 10’’, 65°C for 30’’, 72°C for 30’’, and a final extension step at 72°C for 5’ and stored at 4°C.

### Library preparation and Sequencing

For all libraries, each sample was pooled (7 μL/sample) and library PCR products were purified with SPRIselect beads (A66514, Beckman Coulter). The PoC, test, and pilot cohorts were purified as follows: ratio 0.8:1 (beads:library), and the extended cohort with 1:1 (beads:library) (Beckman Coulter). Due to NSA products in the fragment analyzer profile (**Extended data Fig. 3c**) in the test cohort and pilot cohort, we performed size selection purification (220-350 bp) using the Pippin Prep system (Pippin HT, Sage Science). Library quality was assessed with the 5200 Agilent Fragment Analyzer (ThermoFisher) and Qubit 2.0 Fluorometer (ThermoFisher). All libraries were sequenced with MiSeq or NextSeq 500 (Illumina) using 75 bp paired-end sequencing.

### COVID-19 (C19-)SPAR-Seq platform

Our Systematic Parallel Analysis of Endogenous RNA Regulation Coupled to Barcode Sequencing (SPAR-Seq) system^8^ was modified to simultaneously monitor COVID-19 viral targets and additional controls by multiplex PCR assays. For barcode sequencing, unique, dual-index C19-SPAR-Seq barcodes were used. Unique reverse 8-nucleotide barcodes were used for each sample, while forward 8-based barcodes were used to mark each half (48) of the samples in 96-well plate to provide additional redundancy. These two sets of barcodes were incorporated into forward and reverse primers, respectively, after the universal adaptor sequences and were added to the amplicons in the second PCR reaction.

### Demultiplexing and Mapping

Illumina MiSeq sequencing data was demultiplexed based on perfect matches to unique combinations of the forward and reverse 8 nucleotide barcodes. Full-length forward and reverse reads were separately aligned to dedicated libraries of expected amplicon sequences using bowtie^23^ with parameters –best -v 3 -k 1 -m 1. Read counts per amplicon were represented as reads per million or absolute read counts. The scripts for these steps are available at https://github.com/UBrau/SPARpipe.

### Filtering of low-input samples

To remove samples with low amplified product, likely reflecting low input due to inefficient sample collection or degradation, before attempting to classify, we computed precision-recall curves for classifying control samples into ‘low amplification’ and ‘high amplification’ based on reads mapped to RNA amplicons but ignoring mapping to genomic sequence, if applicable. The former group comprised all controls in which individual steps were omitted (H2O controls) and the latter comprised HEK293T as well as synthetic SARS-CoV-2 RNA controls. For each PoC, test and pilot runs, we obtained the mapped read threshold associated with the highest F1 score, representing the point with optimal balance of precision and recall. Samples with reads lower than this threshold were removed from subsequent steps.

### SARS-CoV2 positive sample classification

To assign positive and negative samples, we used negative (H2O and HEK293T) and positive (synthetic SARS-CoV-2 RNA dilutions) internal controls for each run and calculated optimum cut-offs for viral reads by PROC which defines the threshold for optimum PPV (positive predictive value) and NPV (negative predictive value) for diagnostic tests. Thus, a sample was labelled positive if it had viral reads above the viral read threshold; negative if it had viral reads below the viral read threshold and human reads above the mapped read threshold; and inconclusive if it had both viral and human reads below the respective thresholds.

### Sample classification by heatmap clustering

Heatmap and hierarchical clustering of viral and control amplicons, log_10_(mapped reads+1), was used to analyze and classify all samples. Samples with a total mapped read count lower than the RNA QC threshold were labeled as inconclusive and removed before the analysis. Known positive (high, medium, and low) and negative control samples were used as references to distinguish different clusters. In addition, dilutions of synthetic SARS-CoV-2 RNA were also included as controls and analyzed across different PCR cycle and primer pool conditions.

### Viral mutation assessment

To remove PCR and sequencing errors for the assessment of viral sequence variations, we determined the top enriched amplicon sequence. For this, firstly, paired end reads were stitched together to evaluate full length amplicons. The last 12 nucleotides of read1 sequence are used to join the reverse complement of read2 sequences. No mismatches were allowed for stitching criteria. The number of full length reads per unique sequence variation were counted for each amplicon per sample by matching the 10 nucleotides from the 3’ and 5’ end of the sequence with gene-specific primers. (scripts are available at https://github.com/seda-barutcu/FASTQstitch and https://github.com/seda-barutcu/MultiplexedPCR-DeepSequence-Analysis) The top enriched sequence variant from each sample is used for multiple alignment analysis using CLUSTALW.

### Non-specific amplicon assessment

Single-end reads that contain the first 10 nucleotides of the illumina adaptor sequence were counted and binned into relevant forward and reverse gene specific primer pools by matching the first 10 nt of the reads with primer sequences. Relative abundance of the non-specific amplicons was quantified as percentage of the reads corresponding to non-specific amplicon per forward or reverse primer (Scripts are available at https://github.com/seda-barutcu/MultiplexedPCR-DeepSequence-Analysis).

### Data Availability

Data submitted to GEO (accession number pending).

### Code Availability

We provided the code for demultiplexing and mapping at https://github.com/UBrau/SPARpipe, viral mutation assessment and non-specific amplicon assessment at https://github.com/seda-barutcu/FASTQstitch https://github.com/seda-barutcu/MultiplexedPCR-DeepSequence-Analysis.

## Data Availability

All data-sets and corresponding FASTQ files were submitted to GEO (accession number pending).

## Acknowledgments

The authors thank Drs. Rita Kandel and Jim Woodgett for discussions. The authors are grateful to Tanja Durbic and Kyle Turner in the Donnelly Sequencing Center for sequencing samples and Kathy Fung at the Network Biology Collaboration Center.

## Funding

This work is supported by the Toronto COVID-19 Action Initiative (TCAI) Fund from the University of Toronto awarded to J.L.W, L.P. and R.B., and by a donation from the Krembil Foundation (SHSF Krembil SARS-COV-2) to J.L.W, L.P. and R.B.

## Author contributions

J.L.W., M.M.A, J.J.H., H.H., U.B., B.B. and S.B. designed the study. M.M.A, J.J.H. performed C19-SPAR-Seq experiments. S.B. performed non-specific amplicon/mutation analysis, S.B. and U.B. performed NGS analysis and established C19-SparSeq interpretation pipeline, H.H. performed unsupervised clustering, M.M.A. and J.J.H. assisted with the rest of the analysis. D.T. collected the samples and purified the RNA. J.P.D. performed qPCR control studies (‘BGI’) under supervision of R.B. K.C. performed sequencing. M.B. and M.J. assisted with automation optimization. S.L.P., K.J., and S.S. prepared control samples under supervision of L.P. and L.A.. T.M., S.P, C.B., and B.H. provided access to patient samples, collection of diagnostics information, and assembly of the cohorts. All experiments were carried out under the supervision of B.B, L.P and J.L.W. The manuscript was written by M.M.A., J.J.H., S.B., U.B., L.P. and J.L.W. with input from R.B., T.M., S.P., L.A., H.H., and B.B..

## Ethical declarations

### Competing Interests statement

J. Wrana is founder and CEO of iTP Biomedica Inc, which employs whole transcriptome NGS tests in cancer, and he is founder and consultant for Fibrocor LP, which is developing therapeutics for fibrotic disease. The other authors declare no competing interests.

## Extended data Figures

**Extended data Fig. 1:**
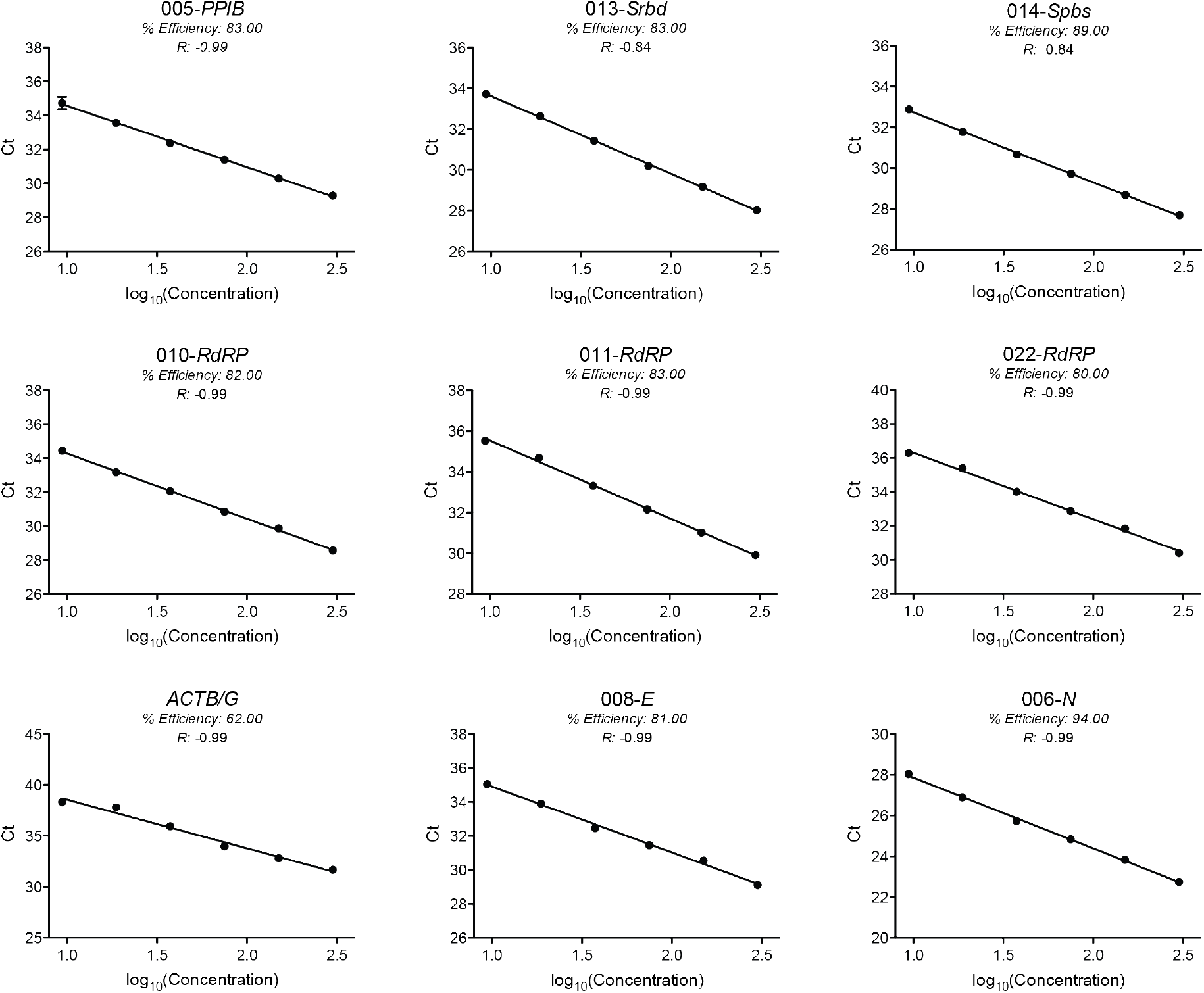
Efficiency of multiplex primers. Standard curve of Ct values (Y-axis) and log_10_(Concentration) (X-axis) of 6 limited dilutions of SARS-CoV-2^high^ sample (LTRI-18) for 9 pairs of primers (see Supplementary Table S1). Each condition was tested in duplicate. Means are plotted for each point. The percent efficiency and the correlation (r) are calculated for each pair of primers after linear regression.

**Extended data Fig. 2:**
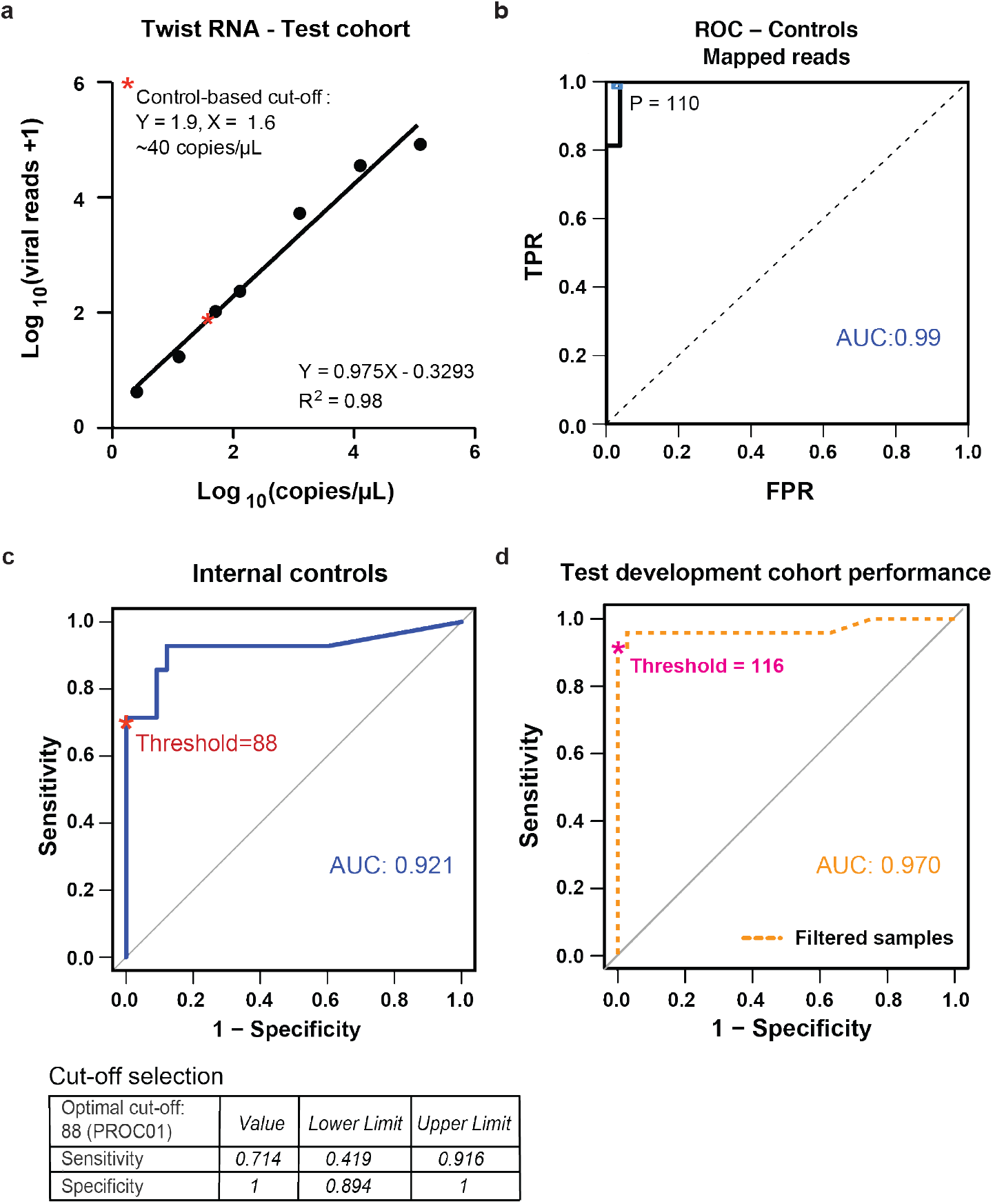
Using embedded controls as a training set for a control-based PR and ROC classifier. **a**, Total viral read counts are plotted against estimated viral copies (copies/μL) obtained using synthetic Twist SARS-CoV-2 RNA with statistics indicated. The cutoff defined by PROC analysis (see panel **c**) is marked with a red asterisk. **b**, Thresholding sample quality. coPR analysis on control samples: ROC of control samples for accurate detection of mapped reads are plotted. The optimal precision and recall read cut-off associated (P = 110) with the highest F1 (0.97) score, and AUC (area under the curve) is indicated on the ROC plot. **c**, Threshold for classification of positives in the test cohort. Total viral reads of negative (H2O and HEK293T) and positive (Twist dilutions) samples are used to calculate optimum cut-off by PROC and the defined threshold (P = 88) is plotted on the ROC curve. Values of sensitivity, and specificity at this cut-off are indicated (below). **d**, Performance of C19-SPAR-Seq. ROC analysis on patient samples that passed RNA-QC threshold was performed using clinical diagnostic results (Seegene Allplex qRT-PCR assay, **Supplementary Table 3**) and total viral reads for patient samples (n = 112). AUC is indicated on the graph.

**Extended data Fig. 3:**
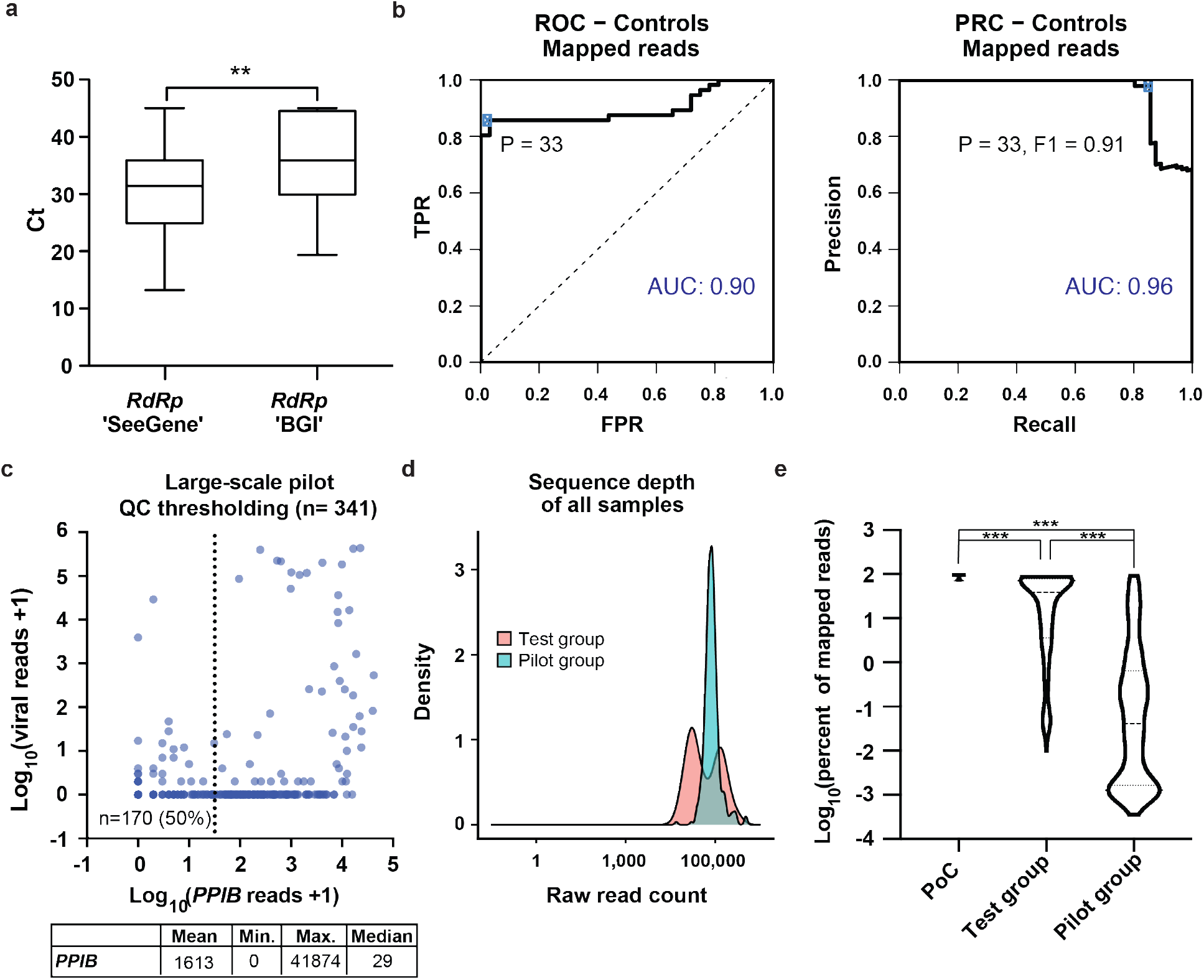
Quality metrics assignment for the pilot cohort. **a**, Comparison of Ct (*RdRP*) values in ‘SeeGene’ *versus* ‘BGI’ tests of the positive archival samples. **b**, coPR analysis on control samples. ROC and PRC of control samples are plotted and the optimal precision and recall cut-off (P = 33) associated with the highest F1 score (0.91) was calculated, as indicated in the PRC plot. **c**, coPR thresholding of the pilot cohort. Plot of total viral reads +1 (Y-axis) *versus PPIB* reads +1 (X-axis) of 341 patient samples in a pilot cohort (see Methods) is shown with the threshold (*PPIB* read counts > 33) to filter low-input samples marked. 170/341 (50%) samples were inconclusive (upper panel). Mean, minimum, maximum, and median values of *PPIB* and total viral read counts are indicated in the table (lower panel). **d**, Sequencing depth of test development and pilot cohort. Distribution density of raw read counts for the test development (pink) and pilot (turquoise) cohorts are shown. **e**, Read mapping percentages. Comparison of overall read mapping percentages between the PoC (**Fig. 1**), test (**Fig. 2**) and pilot cohort (n = 341). One way ANOVA - Tukey’s multiple comparison test (****: *p* < 0.0001).

**Extended data Fig. 4:**
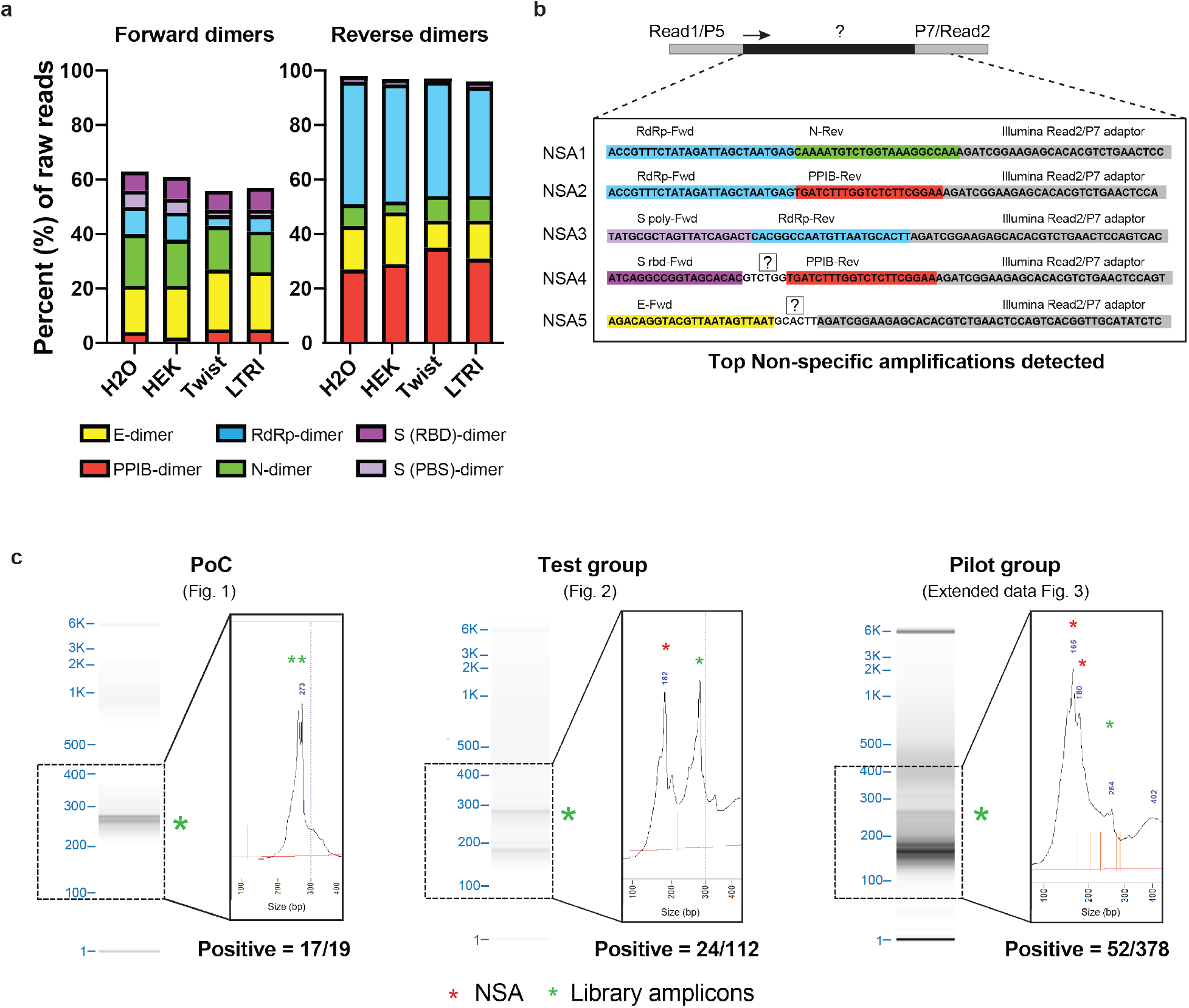
Non-specific amplification (NSA) in pilot cohort. **a**, Analysis of NSAs in the pilot cohort. NSAs contaminating the C19-SPAR-Seq library were quantified and percentage of reads mapping to the indicated forward and reverse primers are plotted. **b**, Schematic examples and sequences of the top 5 NSAs are shown. **c**, Comparison of fragment analyzer profile of the PoC, test development, and pilot cohort libraries after 0.8X SPRI bead purification. Fragment separation (DNA gel) and blow up view of the product abundance (electropherogram) are shown. Expected library amplicons (green stars) and non-specific amplicons (red stars).

**Extended data Fig. 5:**
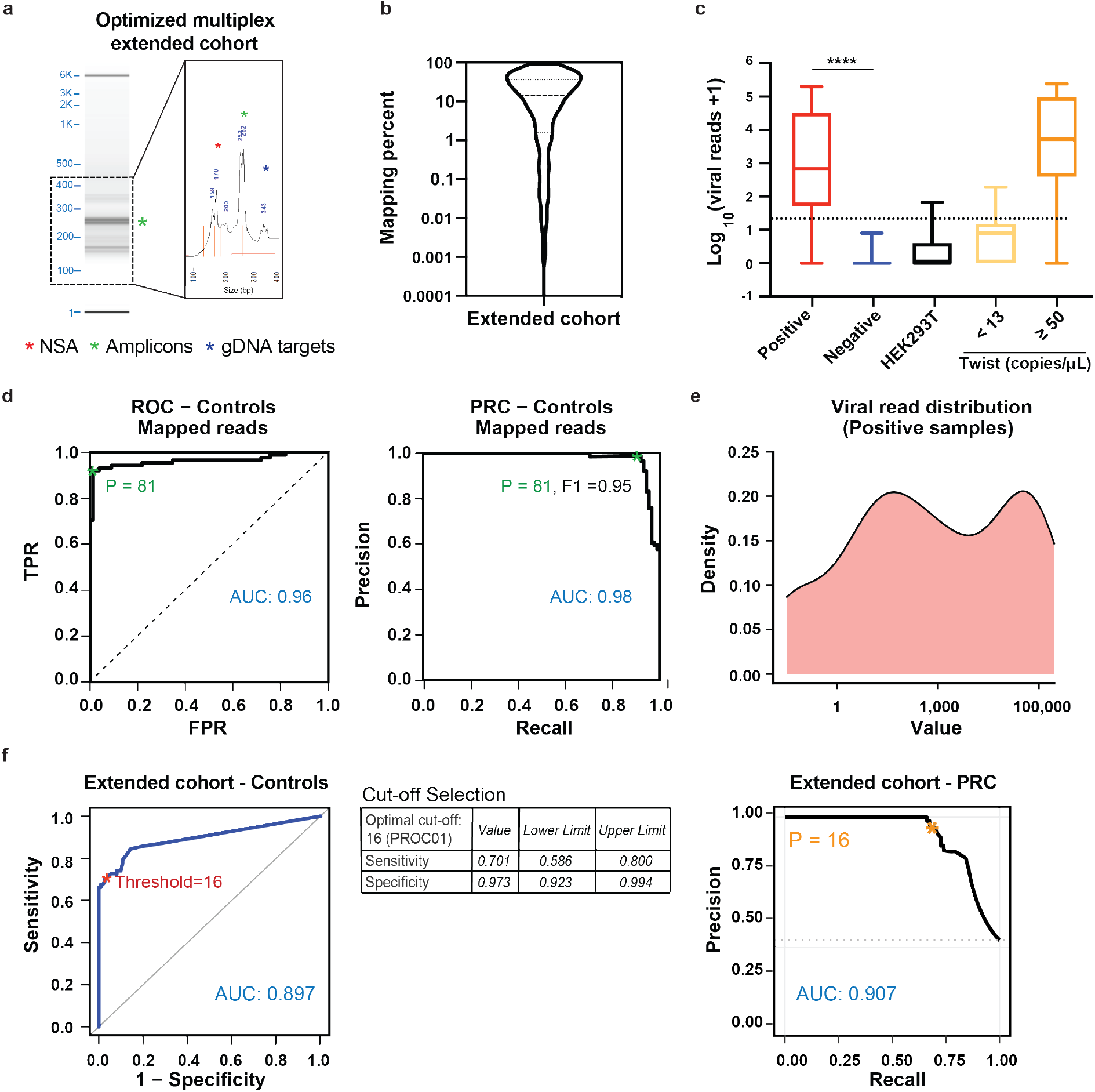
Suppressing non-specific amplicons and quality metrics assignment for the extended cohort. **a**, Fragment analyzer profile of the extended cohort library using an optimized multiplex primer set targeting *ACTB/G, Spoly, Srbd*, and *RdRP*. Fragment separation (DNA gel) and blow up view of the product abundance (electropherogram) is shown. **b**, Mapping percentage of the extended cohort. **c**, Overall distribution of total viral reads in the indicated positive samples (n = 98, red), negative samples (n = 444, blue), HEK293T (n = 21, black), synthetic SARS-CoV-2-RNA (< 13.2 copies/μL, n = 6, yellow), and synthetic SARS-CoV-2-RNA (>= 50 copies/μL, n = 30, orange) are plotted. Unpaired *t*-test of negative *versus* positive samples (****: *p* < 0.0001). **d**, coPR thresholding of sample quality and classification in the extended cohort. coPR analysis on control samples for sample quality yielded an optimal precision and recall read cut-off (P = 81) as indicated. **e**, Distribution of log_10_ total reads +1 of the positive (n = 98) samples. **f**, Threshold for classification of the extended cohort. ROC on control samples (HEK293T and synthetic SARS-CoV-2 RNA control) was assessed to identify an optimal cut-off (P = 16) for classifying patient samples. Performance on the controls is summarized.

**Extended data Fig. 6:**
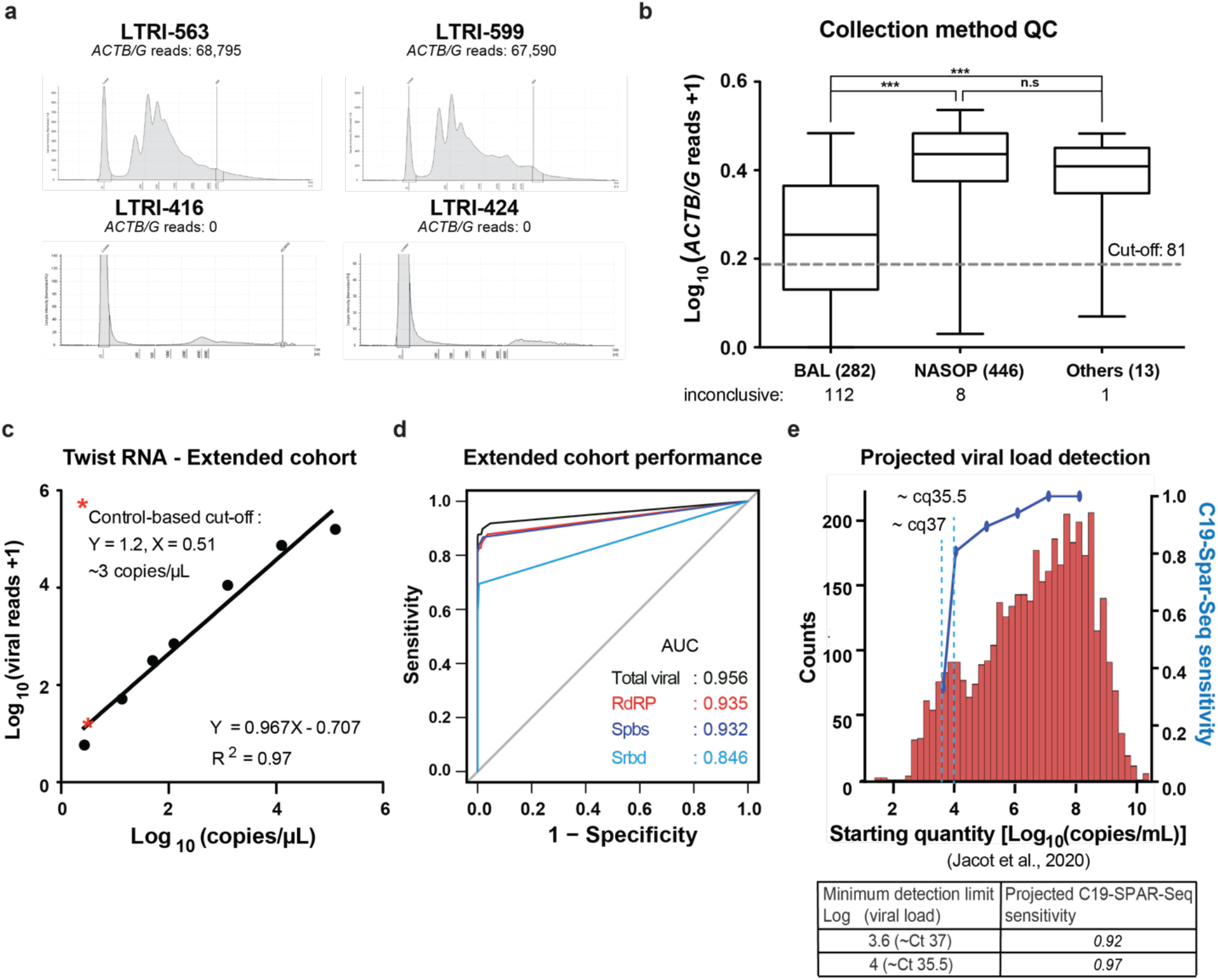
C19-SPAR-Seq performance. **a**, RNA profile of BALs. RNA purified from ten BALs above and 10 below the QC threshold was profiled and two representative traces of each group are shown. *ACTB/G* reads are indicated for each sample. **b**, *ACTB/G* reads according to collection type. *ACTB/G* reads are plotted for each collection type as a box and whisker (median ± 95% confidence interval, and the maximum and minimum values). The number of samples filtered by coPR (ACTB/G reads < 81) are indicated for each group. 1way ANOVA - Tukey’s multiple comparison test (****: *p* < 0.0001, ns: non significative) **c**, Standard curve of total viral reads plotted against synthetic SARS-CoV-2 RNA concentrations obtained from C19-SPAR-Seq analysis of the extended cohort. **d**, ROC curve analysis was performed for each of the indicated viral amplicons and the AUC is shown. **e**, Projection of our C19-SPAR-Seq sensitivity onto the viral load data of ∼4,000 patients from Jacot *et al*., 2020 study^19^. Minimum detection limit and C19-SPAR-Seq sensitivity values are indicated in the table below.

## Supplementary information

**Supplementary Table 1: List of SARS-CoV-2 and human primers**. Primer sequences, name of the targeted regions, size of amplicons after multiplex and barcode PCR are indicated.

**Supplementary Table 2: Itemized cost of C19-SPAR-Sseq per sample**

**Supplementary Table 3: Description of the proof-of-concept cohort for C19-SPAR-Seq detection of SARS-CoV-2**. Barcodes ID, sample identification (ID), date of retrieval, collection method, diagnostic laboratory status, and ‘BGI’ qRT-PCR results are indicated. These patient samples were used to develop C19-SPAR-Seq detection of SARS-CoV-2 (PoC cohort) (**Fig. 1**).

**Supplementary Table 4: Description of test development cohort**. Barcodes ID, sample identification (ID), date of retrieval, collection method, diagnostic laboratory qRT-

PCR results (‘Seegene’) are indicated (n = 112). These patient samples were used to establish SARS-CoV-2 clinical status assignment using diagnostic laboratory qRT-PCR results (‘Seegene’) and to test C19-SPAR-Seq detection of SARS-CoV-2 (**Fig. 2**,**3**).

**Supplementary Table 5: Confusion matrix of the test development cohort**.

**Supplementary Table 6: Description of the pilot cohort**. Barcodes ID, sample identification (ID), date of retrieval, collection method, diagnostic laboratory qRT-PCR results (‘Seegene’), ‘BGI’ qRT-PCR results are indicated. Filtered archival samples are indicated. (Extended data **Fig. 3**,**4**).

**Supplementary Table 7: Description of the extended cohort**. Barcodes ID, sample identification (ID), date of retrieval, collection method, diagnostic laboratory qRT-PCR results (‘Seegene’), and ‘BGI’ qRT-PCR results are indicated (**Fig. 4**).

**Supplementary Table 8: Confusion matrix of the extended cohort**.

**Supplementary Table 9: Group classifications**

## Notes

### Author Declarations

Mount Sinai Hospital Research Ethics Board (Study #20-0078-E)

